# Longitudinal SARS-CoV-2 infection study in a German medical school

**DOI:** 10.1101/2021.05.04.21256382

**Authors:** Michael Schön, Clemens Lindenau, Anja Böckers, Claire-Marie Altrock, David A. C. Messerer, Lydia Krys, Anastasia Nosanova, Nicole Lang, Andrea Renz, Joris Kroschel, Alexandra Beil, Elke Pensel, Claudia Grab, Benjamin Mayer, Ulrich Fassnacht, Jan Philipp Delling, Magdalena Engelmann, Astrid Horneffer, Maria Zernickel, Klaus-Michael Debatin, Jan Münch, Frank Kirchhoff, Thomas Wirth, Tobias M. Boeckers

## Abstract

In light of the COVID-19 pandemic, universities around the world were challenged by the difficult decision whether classes could be held face-to-face in the winter semester 20/21. The gross anatomy course is considered an essential practical element of medical school. In order to protect the participants and teaching staff and to gain more knowledge about SARS-CoV-2 infections among students during a semester with face-to-face teaching a longitudinal test study was conducted. Medical students from the first three years of medical school were also invited. Out of a total of almost 1,000 swabs, only two active asymptomatic infections were detected at the start of the semester, none during the semester. At semester start, approximately 6% of the students had antibodies. At the end of the semester, only nine seroconversions after infection in 671 individuals occurred. This was surprisingly low because a massive second wave of infections hit Germany during the same period. The conclusion therefore is that face-to-face teaching under these measures was not infection-promoting even with high incidence rates in the overall population with the SARS-CoV-2 variants present at that time period. Moreover, the results are indicative of a preventive effect of hygiene concepts together with repetitive testings before and during a semester.

## 1. Introduction

Infection with SARS-CoV-2 is associated with mild to severe acute respiratory symptoms. In rare cases, the infection is asymptomatic. Particularly in severe disease courses, multiorgan disease may occur^1,2,3,4,5^. Young, healthy individuals show usually milder symptom than elderly individuals^2,4,6^. Susceptibility to infection, however, increases sharply in the age range between 18 and 25 years^6^. The outbreak of the pandemic in spring 2020 led to discontinued or reduced classroom teaching at schools and universities, including medical schools worldwide as well as in Germany^7^. For the past winter semester (at Ulm University from November 2020 to March 2021), extensive hygiene concepts were developed at Ulm University, as well as at many other universities worldwide, in order to be able to hold essential practical courses as conventional classroom teaching. In early fall 2020, mainly younger people were infected (Robert Koch Institute, daily situation report, 20 Oct 2020, in German: https://www.rki.de/DE/Content/InfAZ/N/Neuartiges_Coronavirus/Situationsberichte/Okt_2020/2020-10-20-de.pdf?__blob=publicationFile), which was associated with a subsequent second infection wave in Germany. In view of the sharp rise in infection numbers in fall, which indicated the onset of a severe second infection wave in Germany, we developed a longitudinal testing strategy with direct pathogen detection in addition to a hygiene concept already developed in spring and summer for the macroscopic (gross) anatomy course at Ulm University and further semester cohorts from the first three years of medical and dental school. This course is considered essential for medical school training^8^. In order to gain more information on the incidence of infections in students, this study was supplemented with antibody detections at the beginning and end of the semester. See a full definition of the aims of the study in **Supplementary Table 1**. A similar approach has already been proposed by American colleagues^9^. In a modeling of infection events, one to two tests with direct pathogen detection in students before and during a semester were also recommended^10^.

## 2. Design and methods of the study, hygiene concept

All students of the first three years of medical school (semesters 1, 3, 5, and 6), as well as dental students of the first and second semester were invited to participate voluntarily after informed written consent. In Germany, medical school lasts six years and is made up of a preclinical (first two years), clinical-theoretical (third year), and clinical phase. The core cohort of the study was the gross anatomy course with students and institute staff. Cohort sizes, participation rates with corresponding average length of practical courses with face-to-face teaching can be seen in **Table 1** and in **Supplementary Figure 1** in the appendix.

**Table 1:** Cohorts, participants, and amount of face-to-face teaching The time data for face-to-face teaching apply to most students in the cohorts. Exams in classroom are not indicated. A person has participated per definition if he or she attended at least one of the antibody tests. *Times of conventional classroom teaching for the anatomical anatomy course have been adopted. Note that tutors had several other face-to-face classes depending on semester affiliation (mostly semester 5), which are not listed here. University lecturers partly taught in one (84 TU) or two courses (168 TU). Numbers after D and HM indicate the semester affiliation. AC = gross anatomy course, D = dentistry, f2f = face-to-face, HM = human medicine, TU = teaching unit (1 TU equivalent to 45 min).

|  | **n (cohorts invited)** | **n (participants)** | **face-to-face teaching (TU)** |
| --- | --- | --- | --- |
| **all** | **1103** | **868** |  |
| AC students | 385 | 362 | 84 |
| AC tutors* | 66 | 66 | 84 |
| AC teaching staff* | 16 | 14 | 84-168 |
| HM1 | 344 | 231 | 58 |
| D1 | 24 | 15 | 6 |
| D2 | 24 | 13 | 211 |
| HM5 | 185 | 137 | 63 |
| HM6 | 51 | 24 | 76 |
| no f2f teaching | 8 | 6 | 0 |

Testing consisted of direct pathogen detection (pharyngeal swab with reverse transcription polymerase chain reaction (RT-PCR) analysis, rapid antigen test as self-testing) and serum antibody tests. The basic study design is outlined in **Figure 1**. The swab tests with RT-PCR analysis were offered at neuralgic time points with an increased risk of infection: before the start of the gross anatomy course (early November) and after the Christmas break (early January). At both time points, many participants spent the time before in their home environment. To protect the relatives and contacts of the study participants at home, we also offered rapid testing before Christmas break and after the end of the semester. To assess infection prevalence before and during the semester, serum antibodies to SARS-CoV-2 at the beginning of the semester (mid-November to early December) and at the end of the semester (mid-February to early March; approximately three months after the first testing time point) were determined. A paper-based questionnaire with documentation of age, sex, and vaccination status accompanied these two antibody testings. Students in the first-year cohorts (HM1, D1, and D2; see **Table 1**) received all antibody tests, but only the offer of a swab RT-PCR test at the beginning of the semester (late November) and no rapid tests. These cohorts mostly had few courses with conventional classroom teaching at the university. The same applies to fifth- and sixth-semester students (HM5 and HM6; see **Table 1**), who also did not receive swab RT-PCR testing. Most of these third-year cohorts had already been tested for SARS-CoV-2 infection with a swab outside of this study due to clinical courses. The cohort of the gross anatomy course was offered direct pathogen detection more frequently (second swab with RT-PCR, two antigen rapid tests) because of the long duration of the face-to-face classes and special conditions. The dissection of a body donor, even with a reduced number of students, requires an undercutting of the recommended spatial distance (see hygiene concept of the course of macroscopic anatomy in the appendix, **Supplementary Table 2**).

**Figure 1:**
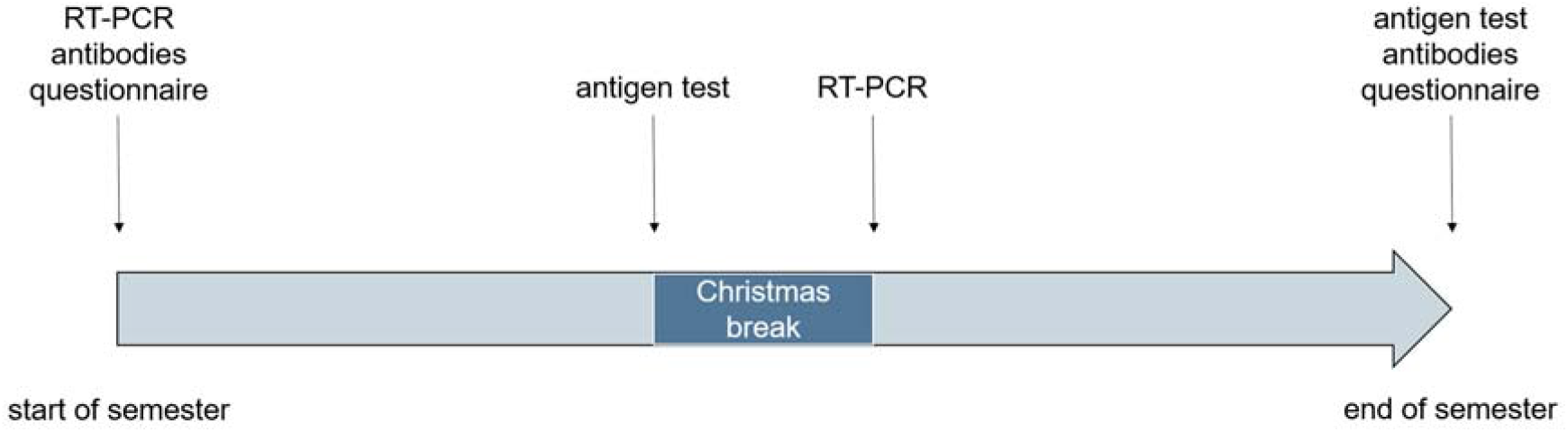
Study design This testing offer was made to all anatomy course students, student tutors, and staff. The other semester cohorts were invited to the antibody determinations at the beginning (mid-November to early December 2020) and end of the semester (mid-February to early March 2021). In some cohorts, a swab test was offered at the beginning of the semester (HM1, D1, and D2).

In the case of underage persons (n=4), a swab test was offered and only executed by a study physician, but the result was not included in the data analysis.

The data from the tests were then calculated as percentages and for the antibody tests corrected according to sensitivity and specificity.

### Hygiene concept in the gross anatomy course

The hygiene concept for the gross anatomy course is summarized as an example in the appendix (**Supplementary Table 2**). Central elements of the hygiene concept were contact tracing, reduction of group size per body donor (six students, one tutor; note: before pandemic ten to eleven students), prohibition of contact between groups, spatial distance (> 1.5 m where possible), mandatory wearing of masks (surgical or FFP), high air exchange rate, disinfection of teaching materials, and regular informational offerings during course time and via e-mails or as online teaching events. There was a maximum of 116 persons in total with teaching staff at the same time in dissection rooms covering 450 m^2^. Within the table groups, it was attempted to maintain as much distance as possible, but it was usually less than 1.5 m.

### Pharyngeal swab with SARS-CoV-2 RT-PCR analysis

A naso- and oropharyngeal swab (same swab) was performed. At the request of the participants, in some cases only an oropharyngeal swab was preferred. Exclusion criteria were symptoms suspicious for COVID-19, as these prohibited entry into the university (see first chapter in the results section, second paragraph). Senior medical students in the clinical phase were appropriately trained in the performance of this testing and instructed in essential hygiene measures. A dry swab from the company nerbe plus (Winsen, Germany) was used. The analysis was performed using the Cobas 6800 test system from Roche Diagnostics (Mannheim, Germany). According to the manufacturer, sensitivity and specificity are both at 100% (correspondence with Roche Diagnostics, April 2021). Ten swab sticks were pooled for each test according to the multiple swab method^11^. The swabs were retained until a negative pool result was obtained; if the result was positive, the pool was dissolved. Roche Cobas gives positive or negative results for two different target regions: *ORF1 a/b* (non-structural region unique to SARS-CoV-2) and the *E* gene (envelope structural protein used to detect pan-sarbecoviruses, but also used to detect SARS-CoV-2 virus).

### Antigen Point-of-care (POC) test

The SARS-CoV-2 Rapid Ag from Roche Diagnostics (Mannheim, Germany) test was used. The test systems were separated and packed in sterile bags and offered for takeaway. Sensitivity and specificity is 99.03 and 98.65%, respectively (correspondence with Roche Diagnostics, April 2021). For name-related data protection and hygiene reasons, this test was offered as a self-test. On a defined day, the participants were asked to perform the test on themselves. A self-made, detailed demonstration video and written instructions were provided. If the test result was unclear or positive, students could contact a study physician. This physician performed another antigen test at their home on the same day. A second swab with RT-PCR analysis was also performed.

### SARS-CoV-2 antibodies

Blood was collected using a butterfly collection system and a serum monovette (SARSTEDT, Nümbrecht, Germany). Senior medical students with clinical experience were instructed in the technique and hygiene measures. Elecsys Anti-SARS-CoV-2 S from Roche Diagnostics (Mannheim, Germany) was used as test system. Serum was collected by centrifugation and immunological *in vitro* detection of antibodies (according to manufacturer’s information on request mainly IgG, but also IgM and IgA) against the spike (S) protein receptor binding domain (RBD) of SARS-CoV-2 was performed. The test has been validated on a large collective with representative patient groups and, according to the manufacturer, achieves a sensitivity of 98.80% and specificity of 99.98% (correspondence with Roche Diagnostics, April 2021). An online calculation tool was used to calculate the corrected prevalence, taking into account sensitivity, specificity, and number of individuals analyzed: https://epitools.ausvet.com.au/trueprevalence (accessed date: 20.04.2021). The confidence interval (Blaker) was set at 0.95 for the corrected prevalence.

### Questionnaire

A paper-based questionnaire (format: Evasys v8.1, evasys GmbH, Lüneburg, Germany) was offered at the beginning and end of each semester at the same times as the tests took place. Age, gender and vaccination status against SARS-CoV-2 was asked.

### Data maintenance, information services, and student support

To comply with data protection regulation, all data were pseudonymized by the Center for Clinical Studies, Ulm University Hospital, and were only accessible to the study team in this form. Individual results were communicated via this office.

Information was provided to raise students’ awareness of the pandemic and infection prevention and control measures in the context of their medical courses. At the beginning of each day in the gross anatomy course, information on hygiene measures was communicated according to the current situation. The participants were regularly informed about the interim results of the study, scientific-medical background of the pandemic and the tests performed. For the gross anatomy course students and tutors, e.g., the anatomical conditions of the pharyngeal swab examination with swab sticks were demonstrated on a midsagittal section of a body donor’s head at the tables and could be tried out by the students themselves. The different detection methods of SARS-CoV-2 antibodies, RT-PCR analysis, and the antigen test were shown on graphs (see **Supplementary Figure 2**) in addition to a self-produced video on how to perform the rapid test.

Due to the situation at Christmas and to protect the home environment of the students and staff, three face-to-face course days were canceled and held as an online event. That coincided with a nationwide lockdown. The hygiene concept was continuously adapted to the current requirements of state and university regulations and communicated to the students.

All participants were offered a compensation option if they missed a course as a result of isolation due to a positive pathogen detection. We also provided support to participants if they tested positive outside of the study. For example, we swabbed after the end of the official isolation period (authors’ note: e.g., ten days in case of an asymptomatic course), as a negative test result after given time intervals was a prerequisite by university for resuming courses with face-to-face instruction.

At any point, participants could join or withdraw from the study. In particular, swab and rapid antigen tests were also offered for non-study participants after informed written consent for the test.

### Data availability statement

Data not included in the manuscript may be released upon reasonable request to the corresponding authors and in compliance with the ethics vote.

## 3. Results

The results of all participants are reported in **Table 2** and **Figure 2** (swab RT-PCR), and **Table 3** and **Figure 3**, and **Supplementary Figure 3** in the appendix (serum antibodies). No participant had a medical condition that would have precluded participation in the study. No complications due to testing occurred.

**Table 2:** Results of pharyngeal swabs with RT-PCR among the cohorts D = dentistry, f2f = face-to-face, HM = human medicine, T1= swab start semester, T2= swab after Christmas break.

|  | **n (swab RT-PCR positive T1)** | **%** | **n (swab RT-PCR positive T2)** |
| --- | --- | --- | --- |
| **anatomy course** | 2 out of 402 | 0.5 | 0 out of 429 |
| students | 2 out of 327 | 0.6 | 0 out of 353 |
| tutors | 0 out of 60 | 0 | 0 out of 62 |
| teaching staff | 0 out of 15 | 0 | 0 out of 14 |
| **HM1** | 0 out of 151 | 0 | not offered |
| **D1** | 0 out of 5 | 0 | not offered |
| **D2** | 0 out of 8 | 0 | not offered |
| **HM5** | not offered |  | not offered |
| **HM6** | not offered |  | not offered |
| **no f2f teaching** | 0 out of 2 | 0 | not offered |
| **total** | 2 out of 568 | 0.4 | 0 out of 429 |

**Table 3:** Results of the antibody determination Corrected prevalences in parentheses in the columns with percentages. The percentages after T2 were calculated as follows: seropositives - seropositives after vaccination / all participants at this testing round - seropositives after vaccination. Ab = SARS-CoV-2 antibody, D = dentistry, f2f = face-to-face, HM = human medicine, T1 = antibody test semester start, T2 = antibody test semester end, vacc. = vaccination.

|  | *n (ab test positive T1)* | *%* | *n (ab test positive T2 (seropositive after vacc.))* | *% without seropositives after vacc.* | *n (seroconversion without vacc.)* |
| --- | --- | --- | --- | --- | --- |
| ***anatomy course*** | 23 out of 417 | 5.5 (5.6) | 40 out of 422 (14) | 6.4 (6.4) | 2 |
| *students* | 22 out of 345 | 6.4 (6.4) | 35 out of 342 (10) | 7.5 (7.6) | 2 |
| *tutors* | 1 out of 61 | 1.6 (1.6) | 5 out of 66 (4) | 1.6 (1.5) | 0 |
| *teaching staff* | 0 out of 11 | 0 | 0 out of 14 | 0 | 0 |
| ***HM1*** | 9 out of 155 | 5.8 (5.9) | 38 out of 213 (20) | 9.3 (9.4) | 5 |
| ***D1*** | 0 out of 5 | 0 | 0 out of 14 | 0 | 0 |
| ***D2*** | 2 out of 8 | 25 (25.3) | 2 out of 12 | 16.7 (16.9) | 0 |
| ***HM5*** | 10 out of 111 | 9 (9.1) | 35 out of 132 (18) | 14.9 (15.1) | 1 |
| ***HM6*** | 2 out of 20 | 10 (10.1) | 3 out of 21 (1) | 10 (10.1) | 0 |
| ***no f2f teaching*** | 0 out of 4 | 0 | 1 out of 4 | 25 (25.3) | 1 |
| ***total*** | 46 out of 720 | 6.4 (6.5) | 119 out of 818 (53) | 8.6 (8.7) | 9 |

**Figure 2:**
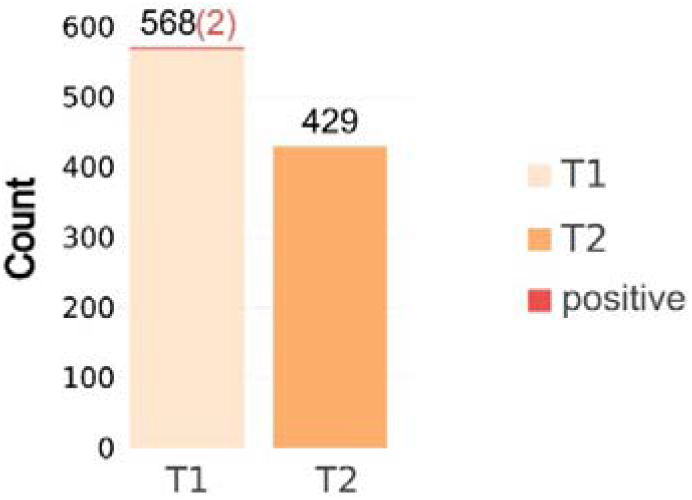
RT-PCR results after swab test T1 = swab start semester, T2 = swab after Christmas break.

**Figure 3:**
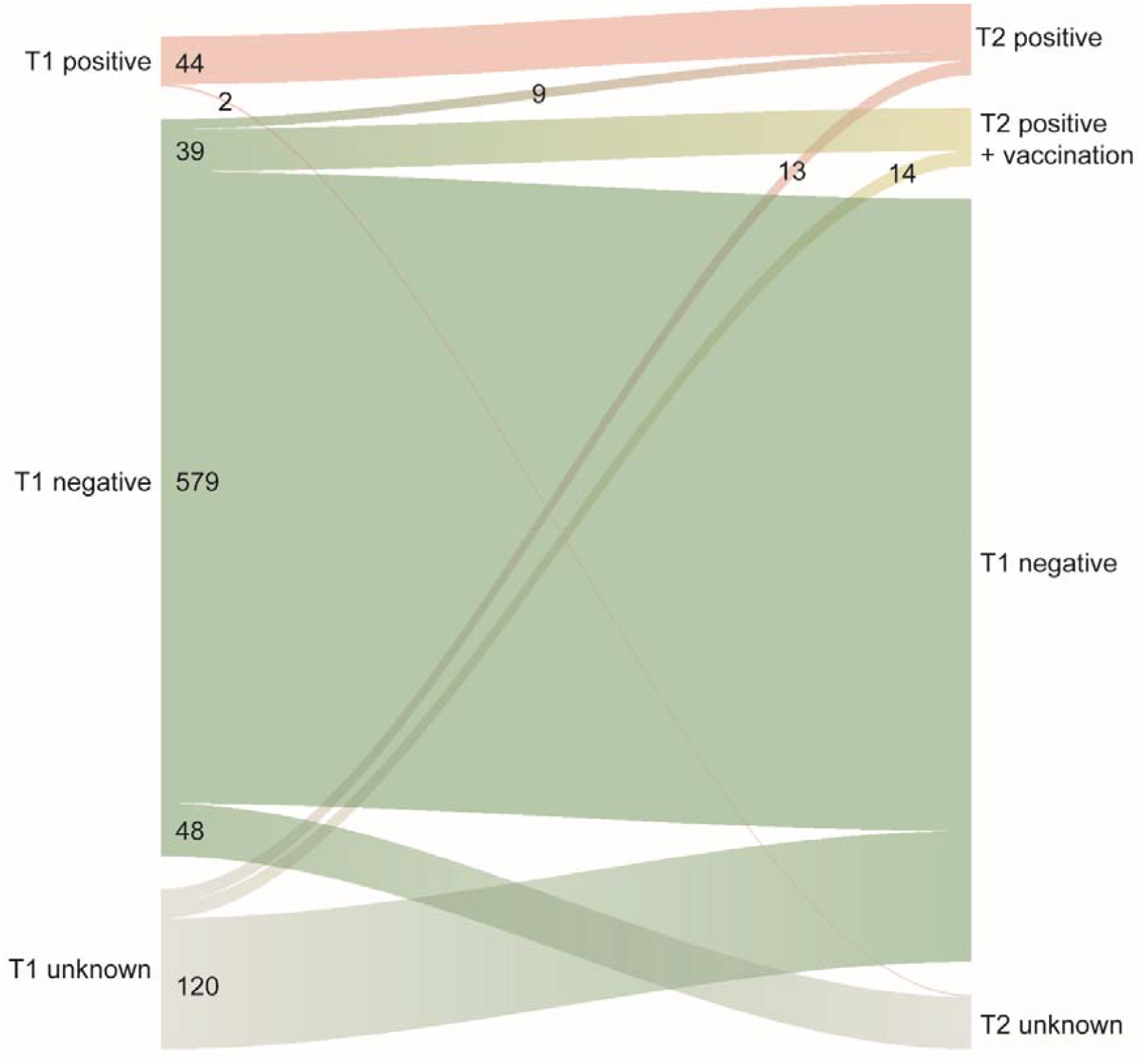
Longitudinal antibody status of all participants The diagram illustrates the antibody status at the start (left, T1) and end (right, T2) of the semester. A negative status is marked green, a positive status after infection is rose-colored, after vaccination yellow. If a participant took part only at T1 or T2, the other test time point is grayed out. Only the green to pink line represents those participants who underwent seroconversion after infection during semester (n=9 of 671 who participated in both tests).

### Description of the study cohorts

A total of 854 students of all invited semester cohorts and 14 staff members of the anatomical institute participated in the serum antibody detections of the study. Participation rates of antibody testing are shown in **Table 1** and **Supplementary Figure 1** in the appendix. They represented nearly 80% of the total number invited participants to the study (n=1103; see **Table 1**). The majority (63.8%) of the participants were female (those invited to the study 61.7%). The mean age was 21.5 years (3.4 years standard deviation) excluding personnel from the anatomical institute. The mean age of all invited students was 21.8 years, standard deviation 3.6 years. The 14 participating institute staff members (n=16) were predominantly university lecturers in the gross anatomy course. The largest invited cohort was the gross anatomy course, which included students (course participants, n=385), student tutors (n=66), and staff (n=16). In this cohort, most participated in both tests (**Supplementary Figure 1** in appendix, see also **Table 1**). Only 6% of the anatomy course students did not participate in any antibody testing. Cohorts in other semesters were smaller and participation rates were lower. The amount of conventional classroom teaching of practical courses can be seen in **Table 1**. Of note, the cohort of the gross anatomy course (students, tutors, staff) had to fall below 1.5 m distance within table groups due to the nature of this course. Therefore, this cohort was offered more tests with direct pathogen detection. The fifth semester study cohort provided most of the student teaching staff (52 of 66 tutors in total) in the gross anatomy course. These are part of the anatomy course cohort for the purposes of this study. Three tutors from semester 6 were included in the anatomy course cohort.

The management of the gross anatomy course was notified of 17 reports of COVID-19 symptoms in course students and tutors during the semester. 15 were quarantined. Nine SARS-CoV-2 infections were documented (three shortly before the semester start, including one tutor), but none could be associated with university teaching events. No contact situation in which a person had to be quarantined as a result of attending the course was reported. There was one SARS-CoV-2 infection in the above-mentioned student tutor in the training week for tutors before the course began. All other tutors as well as the lecturers with contact were classified as category II contacts (authors’ note: low risk of infection; no quarantine necessary) as defined by the Robert Koch Institute. An additionally offered swab with RT-PCR was generally negative. Prior to the swab RT-PCR testing for the anatomy course cohort, six individuals at the start of the semester and five individuals after the Christmas break were unable to participate because they either had a SARS-CoV-2 infection (n=3), symptoms suspicious for COVID-19, or were quarantined.

### Results of the semester start examinations

In the swab test with RT-PCR for the gross anatomy course, 84.9% (n=327) of the course students, 90.9% (n=60) of the tutors, and 93.8% (n=15) of the staff participated. Two swabs of anatomy course students (0.6%) were positive in both target regions, none of the institute and tutor staff. Both positively tested individuals contacted the study team voluntarily and had an asymptomatic infection according to their own statements. We refrain from giving further details on age and sex for reasons of data protection. After ten days, they were offered a swab test with RT-PCR, since a negative test result at this point allowed the resumption of practical courses by the university administration. One individual was then negative. The other individual tested positive in only one of the two target regions (*E* gene) with a now very high Ct (cycle threshold) value (> 34) and was therefore released from quarantine based on the publication by Wölfel et al.^12^. Among first-year human medical and dental students, 164 swabs were negative. All results are provided in **Figure 2** and **Table 2**.

We recorded high participation rates for the first antibody test of the gross anatomy course (see **Table 1** and **Supplementary Figure 1** in the appendix). Twenty-two of the 345 anatomy course students were positive (6.4%), one of them among the tutors (1.6%), and none among institute staff. Participation rates among the other cohorts were lower. Among the 155 tests of human medicine students of the first semester, nine tests were positive (5.8%). Among fifth semester students, ten of 111 tests (9%) were positive. The other cohorts were small and participation rates were not as high as in the larger cohorts. The results are summarized in **Table 3**.

### Results of the second swab with RT-PCR

After the Christmas break, a second swab test was offered before resuming the gross anatomy course. With a participation rate of over 90% among students, tutors, and staff, all 429 samples were negative (see **Figure 2** and **Table 2**).

### Results of the antibody tests at the end of the semester

All results are shown in **Table 3** and **Supplementary Figure 3**. In the gross anatomy course, 35 of the 342 students were detected with antibodies, of which ten vaccinated cases had positive antibody status. 21 of the 22 cases positive at the semester start examination participated again and still had a positive status. Two positive cases with no indication of vaccination were not present at the semester start examination. Thus, only two individuals changed from a negative to a positive finding without having been vaccinated against SARS-CoV-2. Among tutors, five of 66 had a positive status. Of these, four were vaccinated. Because the person who tested positive on the first test participated again and was still positive, we did not see any seroconversions with 100% participation. Among the university lecturers, all 14 were seronegative.

Results among the other cohorts were as follows: Of 213 first-semester human medicine participants, 38 were positive, including 20 after vaccination. Eight of the previously nine positive individuals participated again and were positive again, one of whom was vaccinated.

Five individuals had seroconversion without vaccination, and five seropositive individuals participated only in the second round of testing. Among fifth-semester students, 35 of 132 were positive. Ten of these were so at the semester start test, with one person vaccinated. Thirteen vaccinated individuals were negative at the first test, five had not participated in that first test, and all 18 were positive at the second. Only one person was found to be seroconverted presumably after infection. Six other participants had participated only at the second time point and were positive at that time without vaccination. In the smaller cohorts (dentistry semesters 1 and 2, human medicine semester 6), there were no detected seroconversions despite increased participation rates.

The sex ratio and age distribution of all SARS-CoV-2 antibody-positive individuals (n=66) at semester start or end without vaccination was similar to the overall student study cohort. 42 of the 66 subjects were female, and the mean age was 21.2 years (standard deviation 2.5 years).

A summary over all cohorts in **Supplementary Figure 2** is provided in the appendix. **Figure 3** illustrates the results as flows in the form of a Sankey diagram. It should be noted that some were only present at the start (T1) or end (T2) of the semester (T1 or T2 unknown in **Figure 3**). Especially at the antibody test at semester end, significantly more people participated. Therefore, the percentages after T2, even after removal of those with vaccination, are only comparable to a limited extent with the percentages at the start of the semester (see **Table 3**). It should also be noted that when calculating percentages after T2, those vaccinated were not included.

For a longitudinal comparison, only the individuals who participated at semester start and end (T1+T2) are considered in the analysis of the next paragraph.

### Antibody status of the student cohorts between the start and end of the semester

Overall, we detected only nine seroconversions (+1.36%) among the students who participated in both antibody tests (n=660) that could not be explained by vaccination. **Figure 4** (left pie chart) shows the antibody results of the student participants in the study who appeared at both semester start and end test rounds. 6.7% were already positive at baseline; none had a loss of positive status at semester end. Another nearly 6% became positive after vaccination against SARS-CoV-2. Five vaccinated individuals had a negative antibody result, but of these, either a complete indication of the vaccination date was not available or the first dose was administered only a few days before the test day. As far as indicated in the questionnaire, most of those vaccinated were immunized with the Comirnaty vaccine from Biontech/Pfizer. It is interesting to compare the two larger cohorts of first semester medicine students and the anatomy course students (**Figure 4**; middle and right pie chart, respectively). Note that for the anatomy course nearly 85% of invitees participated in both testing time points, compared to only 40% for semester 1 (see **Supplementary Figure 1**). Although semester 1 had fewer practical courses at university (see **Table 1**), and the gross anatomy course is accompanied by a closer contact between students (within a table group < 1.5 m, see also **Supplementary Table 1** in the appendix), more seroconversions after infection occurred in semester 1.

**Figure 4:**
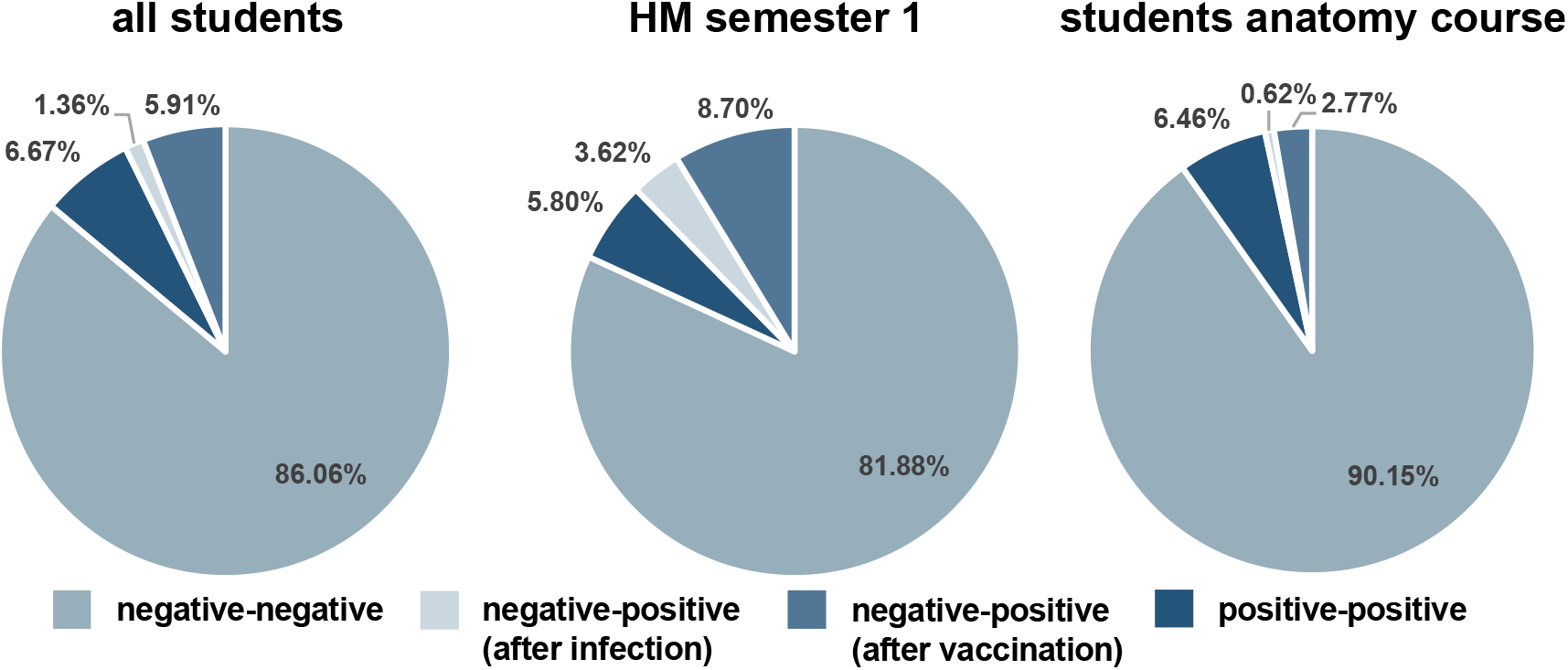
SARS-CoV-2 antibody status in students attending both tests The results refer to those who participated in the tests at the beginning and end of the semester (n=660). Institute staff members (n=11) are not included here. Student tutors are included in the diagram on the left. HM = human medicine.

### Results of the rapid antigen self-tests

A total of four students reported of positive or abnormal test results after both rounds of testing (before Christmas break and end of semester; in total approx. 900 tests). One person had indicated that the swab was very bloody, two had a questionable positive result (very faint or wavy line at T), and one had a positive result. For the last three individuals mentioned, a study physician performed a second rapid antigen test on the same day. The questionably positive tests were now negative, and the swabs with RT-PCR were also negative in each case. The person with a positive antigen test was also positive in the rapid test performed again by the study physician. Pharyngeal swabs with RT-PCR from the same day and two days later were both negative. The study team considered this case as false positive.

## 4. Discussion

### Summary of the study

The study started at the beginning of November and ended beginning of March. Over 60% of students in the anatomy course cohort reported that they felt significantly or very significantly more protected from infection with SARS-CoV-2 as a result of being offered the tests, according to a survey at semester end. In review of the questions asked before the study began in order to protect students and university lecturers and to gain further scientific knowledge (see Supplementary Table 1), important conclusions could be drawn: 1) There were only two asymptomatic infections before the start of the semester, none during. 2) Although more than 6% of students had antibodies to SARS-CoV-2 at the beginning of the semester, there were surprisingly few seroconversions after infections despite a concurrent severe wave of infections in the population. Due to the consistently high participation rate of the anatomy course cohort (including teaching staff) with more than 400 participants in each test (mostly more than 90%), we assume a representative sample of this sub-cohort. A self-selection bias cannot be excluded if, e.g., a person deliberately did not participate after previous infection. However, the infection dynamics of the other semesters between the beginning and end of the semester did not differ fundamentally from that of the anatomy course, although fewer students participated in these groups, and their presence teaching times were also mostly lower. Students in the gross anatomy course actually had fewer seroconversions as result of an infection compared to participants of the other semester cohorts. It was particularly pleasing to note that there was no infection detected by SARS-CoV-2 antibody seroconversion during the semester in the tutors of the gross anatomy course. Most of them belonged to semester 5. In addition to their tutoring activities, they attended the face-to-face practical courses according to their semester affiliation. Among the larger cohorts, only in semester 5 seroprevalence was strikingly higher at the start of the semester. We were informed at the beginning of the semester that a mandatory RT-PCR swab test was performed by the university hospital prior to an examination course in November. Approximately ten out of 100 swabs were positive for SARS-CoV-2. This examination course was then cancelled. The high rate of previously detected SARS-CoV-2 infections most likely explains the higher seroprevalence detected later. This semester was tested for antibodies in early December as part of this study, so this testing most likely covered the post-infectious seroconversions. A limitation of the study is the lack of comparison with a cohort of students who had only online classes during the winter semester.

Many students with little face-to-face teaching at university reported that they would not be living in Ulm, which could explain the lower participation rates at the semester start examinations among these cohorts. At the end of the semester, we were able to pair the timing of blood collection with exams in presence. The steady increase in study participants was encouraging, with a maximum in the last round of testing for antibody determination, demonstrating the high positive acceptance among students. The study team was repeatedly told that the study was very positively received. There was always the possibility to get in contact with the study management to discuss the individual or overall results. This offer was and still is frequently taken. The students showed an extremely high level of interest in protecting themselves and their fellow students, taking active steps against the pandemic such as following infection control measures and participating in the study, and thus ultimately helping to ensure that practical courses can be completed with the necessary presence. Likewise, it was very gratifying and moving for us as a study team to see how many students in the clinical phase were willing to help with the testing rounds. A frequent feedback on their motivation was that they wanted to contribute in dealing with the pandemic and that they could support their preclinical fellow students.

### Interpretation of data in national and international comparison

Few studies have investigated the incidence of SARS-CoV-2 infections in students. In most cases, only seroprevalence at a specific time point was determined^13,14,15,16^. Another study examined antibodies to SARS-CoV-2 in a cohort of students at the beginning of the fall term in the United States. The students had high infection status, with over 30% seropositive findings. The study does not report the antibody status of students at the end of the semester^17^. In the USA, there was a large increase in local COVID-19 incidence in the vicinity of universities that may be associated with the onset of fall term^18^. High seropositivity among students was also reported in medical students from Copenhagen between October and November 2020. About one third of the participants tested positive, which correlated with the attendance of student parties in spring^13^. To our knowledge, a longitudinal study with direct SARS-CoV-2 pathogen and antibody detection during the winter semester is unique. The winter term and the study largely coincided with the second severe wave of the pandemic in Germany (see incidence rates of the city of Ulm and Germany in **Supplementary Figure 4**).

At the start of the semester (testing times from mid-November to early December 2020), 6.4% (46 of 720) of all cohorts were seropositive. This value is difficult to rank in comparison to cohorts of the same age or in an overall population comparison. A survey of students in Los Angeles in May 2020 came up with a value of 4%^14^, and at a Spanish university in July 2020, seroprevalence was less than 3%^15^. However, incidences are not and have not been comparable to other countries. Even within Germany and in each population group, there are likely to be substantial differences. An interim report of the SERODUS I study found a seroprevalence of 3.1% among 18-30 year-old residents of Düsseldorf, Germany. The time of measurement was largely identical to the semester start surveys of this study (https://www.uniklinik-duesseldorf.de/fileadmin/Fuer-Patienten-und-Besucher/Kliniken-Zentren-Institute/Institute/Institut_fuer_Medizinische_Soziologie/Forschung/SeroDus/Feld-_und_Ergebnisbericht_SERODUS-I_SERODUS-II_03-02-2021_v01.pdf, in German). A regionally interesting comparison is a random sample survey of the seroprevalence of the Munich population by the Tropical Institute of the LMU University Hospital Munich in the KoCo19 study. Before Christmas, 3% were seropositive (http://www.klinikum.uni-muenchen.de/Abteilung-fuer-Infektions-und-Tropenmedizin/de/COVID-19/KoCo19/index.html, in German). Like Ulm, Munich is located in Southern Germany. A meta-analysis estimated that between 0.79-3.67% (95% confidence interval) of the population in Germany had SARS-CoV-2 antibodies by August^19^. The SeroTracker program, available online, estimates seroprevalence in nations and regions through a systematic review process. For the testing period (winter term), local prevalence of serum antibodies is estimated to be between 1.4% and 4.4% (https://serotracker.com/en/Explore). In the synopsis of these data, it can be assumed that the values we determined at the start of the semester were above average compared to the German population. This is presumably due to the more frequent social contacts, the housing situation (often shared flats), and the often mild expression of COVID-19 symptoms – or even asymptomatic infections – in this age group^14^. Furthermore, not all individuals develop antibodies after SARS-CoV-2 infection and antibody levels correlate inversely with symptom expression^20^. In addition, antibody levels may decrease over time^21^. Although nearly all seropositive individuals also participated again at the end of the semester three months after the semester start test, not a single test was then negative.

Across all cohorts including institute staff who participated in both antibody tests (n=660 students, n=11 teaching staff), approximately three months after the start of the semester, only nine participants changed from seronegative status to positive at the end of the semester, which cannot be explained by vaccination against SARS-CoV-2. In the gross anatomy course, only two such seroconversions were added to the 21 SARS-CoV-2 seropositive individuals among course students at the start of the semester: an increase of 0.6% in total, and 9.5% when compared to the 6.5% positive individuals at the semester start of those n=325 students who participated in both antibody tests at semester start and end; a 20.5% increase for all n=660 students in this study who participated in both tests. However, in this calculation we do not look at the total study cohort (not everyone participated) and do not consider those who were only present at the test at the start or end of the semester. However, when reviewing percentages after the end of the semester, after exclusion of the vaccinated individuals (see Figure 3), there were no massive increases in prevalence. Thus, we had a strong opposite trend to the infection incidence in Germany. In the same period, the number of all infected persons detected in Germany increased by a approx. 200% (mid-November to end of February, data from the Robert Koch Institute, Germany, and John Hopkins University, USA). Of note, these are not seropositive individuals but persons with a direct detection of the pathogen. However, we assume a similar dynamic between acute infection and the subsequent formation of antibodies. What this opposing trend is due to can only be assumed, especially since clusters of infections among students at the beginning of a semester have been described^21^. It is possible that behavior during the semester break before the winter term differed from that during the semester with regard to disciplined adherence to infection prevention and control measures. The gross anatomy course in particular is considered to be very learning intensive^8,23^. Our impression was that the students felt that it depended on their behavior, individually and on the entire cohort, whether the semester could be carried out as planned. Moreover, we suspect a preventive effect of the tests and the hygiene concepts. The announcement of the tests and the comprehensive information provided about the study and the pandemic likely resulted in a change in behavior towards the pandemic situation.

### Considerations for a testing concept at universities during COVID-19 pandemic

Since costs for testing are likely to play an important role for all university institutions, we recommend selective testing with a swab followed by RT-PCR analysis, especially at the beginning of the semester. The high sensitivity led to the detection of two infected individuals in our study population. It can be derived from our study data that students are more likely to be virus carriers prior to the start of the semester due to returning to the study site from their home environment and lower awareness of the scientific background to the pandemic at the start of the semester. Resource-sparing multiple swab method was sufficient to detect asymptomatically infected individuals in this study. Even then results were available no later than one day after swabbing. Rapid antigen tests were used as self-tests. They are less expensive than RT-PCR testing (authors’ note: approx. ten Euro versus 45 Euro). The rapid antigen test used has high quality in terms of sensitivity and specificity compared to other lateral flow test^24^. According to the study participants, the handling in combination with written and video instructions was sufficient. However, control of use, timing of performance, and feedback of positive results can only be ensured by reliable information from study participants. It may also be that correct performance of the test is more common with knowledge of medical topics. Antibody tests provide a relatively cheap insight into the longer-term infection status of populations. However, we recommend their use primarily for study purposes, as the data are very valuable from a seroepidemiological perspective. It is also possible that feedback on infection incidence by antibody status to students will also generate increased awareness of the pandemic. Further infection epidemiologic studies at other study sites with other collectives would be very important, especially in light of the upcoming more infectious virus variants. Providing regular information on infection prevention and control, scientific and medical background of the pandemic, and testing might also decrease infection numbers due to increased awareness of hygiene recommendations.

We offer to all colleagues in the university environment to support them in the preparation of a study protocol and hygiene concepts, information offers (e.g., video instructions for swab tests, information material on tests), and in the performance of tests on the university campus in compliance with infection prevention and control measures.

Overall, the data of this study indicate that even large face-to-face university teaching classes with more than 100 people and practical courses that require distances between students and teaching staff less than 1.5 m are possible under certain conditions without contributing to an increase in infection rate. The basic prerequisite is a good hygiene concept. Regular provision of information and selective testing presumably lead to preventively effective changes in behavior and might make a decisive contribution to controlling the infection situation. In view of the severity of the pandemic, it cannot be concluded on the basis of the presented data that universities can be opened without restriction. It is necessary to determine which teaching programs are essential to be performed as face-to-face classes. These could then even have an infection-preventing effect with a combination of hygiene concepts, selective testing, and information offers for students. Furthermore, it should be considered whether these measures are sufficient with more infectious SARS-CoV-2 variants, which were not yet strongly prevalent in Germany during the period of the study.

## Data Availability

We would share the study protocol upon request. Release of anonymized data can be discussed in consultation with the Ethics Committee of Ulm University.

## Acknowledgements

The Institute for Anatomy and Cell Biology would like to sincerely thank the following medical students for their voluntary and extraordinary dedication and commitment to sample collection in this study: Josef Anetzberger, Helene Banßhaf, Jonathan Behr, Jacqueline Berg, Amelie Bogenschütz, Carla Brendler, Noemi Damwerth, Osman Demir, Maryse De Molière, Simon Ehricke, Lotta Elonen, Moritz Embacher, Ali Fattom, Oliver Fetzer, Julia Haug, Marlene Heimbeck, Sophie Jauch, Dominik Karl, Bianka Kernl, Tibor Kelety, Tassja Kleiter, Carla Köhler, Anastasia Kosmidou, Sophie Kraft, Pascal Lessing, Lara Linderich, Carolin Lutz, Friederike Lutz, Clemens Miesbichler, Lazaros Milonidis, Adam Mohamed, Antonia Mortsch, Maximilian Münch, Jule Neumann, Elisabeth Oertel, Felix Rabus, Caro Reif, Simon Risel, Leonard Saitta, Franziska Schober, Leon Schurr, Daniel Sommer, Christina Steffke, Johannes Steinhart, Sibylle Steinkellner, Florian Strachwitz-Helmstatt, Elias Stratmann, Laura Stukan, Elisabeth Weissenbach, Nathalie Wimmer, Alexander Ziermann, Leonie Zikarsky, Lena Zöllner. We are grateful to Florian Spuhl and Dr. Roman Wennauer for their competent and reliable help with sample coding and label creation. We thank Katrin Ring and Nadine Pfeifer for skillful laboratory assistance. We thank the Robert Koch Institute for providing study protocols and questionnaire documents^25^. We would like to thank all the participants in the study and the students and tutors of the anatomy course for their exemplary behavior in relation to the pandemic.

## Contributions

MS and TMB designed and led the study. MS wrote the ethical application, study protocol, informed consent form, and hygiene concept. CL and MS organized and conducted the testings. AB, CMA, LK, AN, BM, and MS analyzed the data. NL and AR helped to plan the study and performed name-related care for the data and communicated the individual results to study participants. JK and AlB directed the laboratory analyses of the antibody tests, EP directed the RT-PCR analysis of the pharyngeal swabs. CG, DACM, ME, and AH assisted in organizing the study. UF and AB prepared the hygiene concept for the gross anatomy course. MS provided regular updates to students on the content of the study and the pandemic. JPD assisted with sample collection. MZ and KMD assisted in the preparation of the study protocol^26^. JM and FK advised on virological issues. TW and TMB facilitated funding for the study. MS wrote the manuscript. Major edits were also made by TMB, AB, LK, FK, JM, CMA, and DACM.

## Ethical approval

The study was reviewed and approved by the ethics committee of Ulm University (application number 405/20 and was registered in the German Clinical Trials Register: DRKS00024860).

## Disclaimers

None reported.

## Previous presentations

A previous version of this study has been published as preprint on MedRxiv: https://www.medrxiv.org/content/10.1101/2021.05.04.21256382v2.

---

**Supplementary Figure 1:**
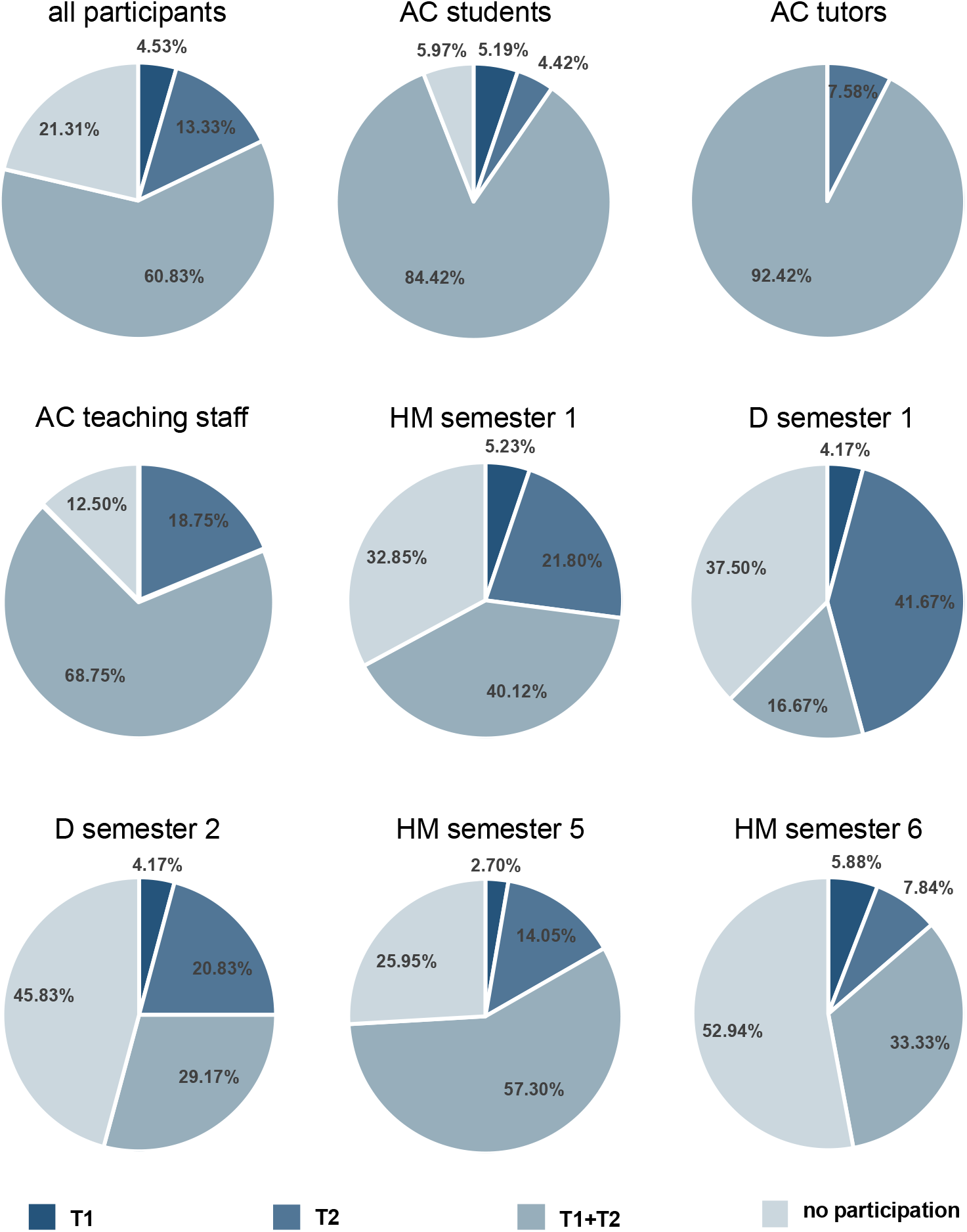
Participation rates for antibody tests in the different cohorts Percentages refer to all those invited for testing (see Table 1). AC = anatomy course, D = dentistry, HM = human medicine, T1 = antibody test semester start, T2 = antibody test semester end.

**Supplementary Figure 2:**
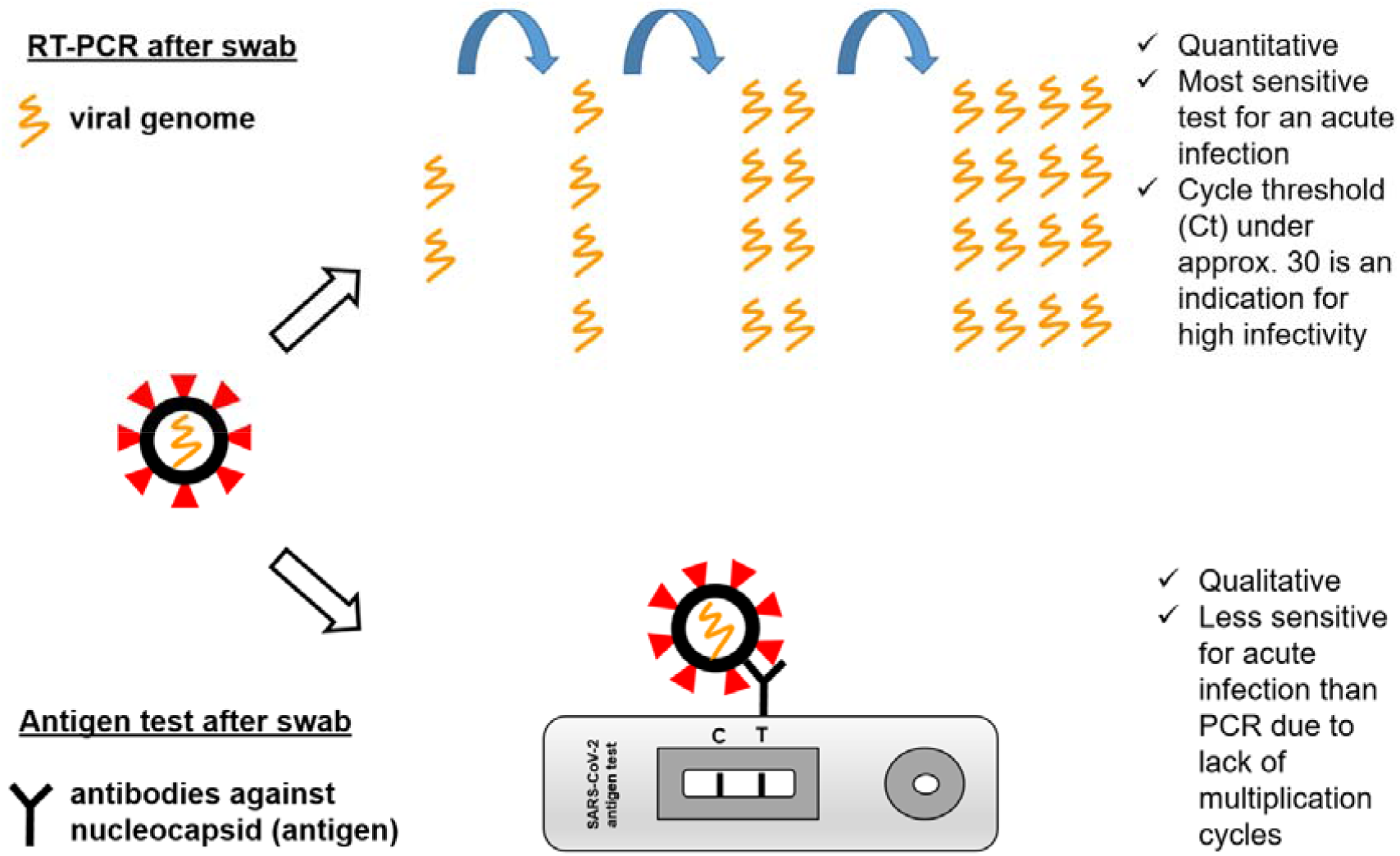
Explanatory graphic for students on how direct pathogen detection works

**Supplementary Figure 3:**
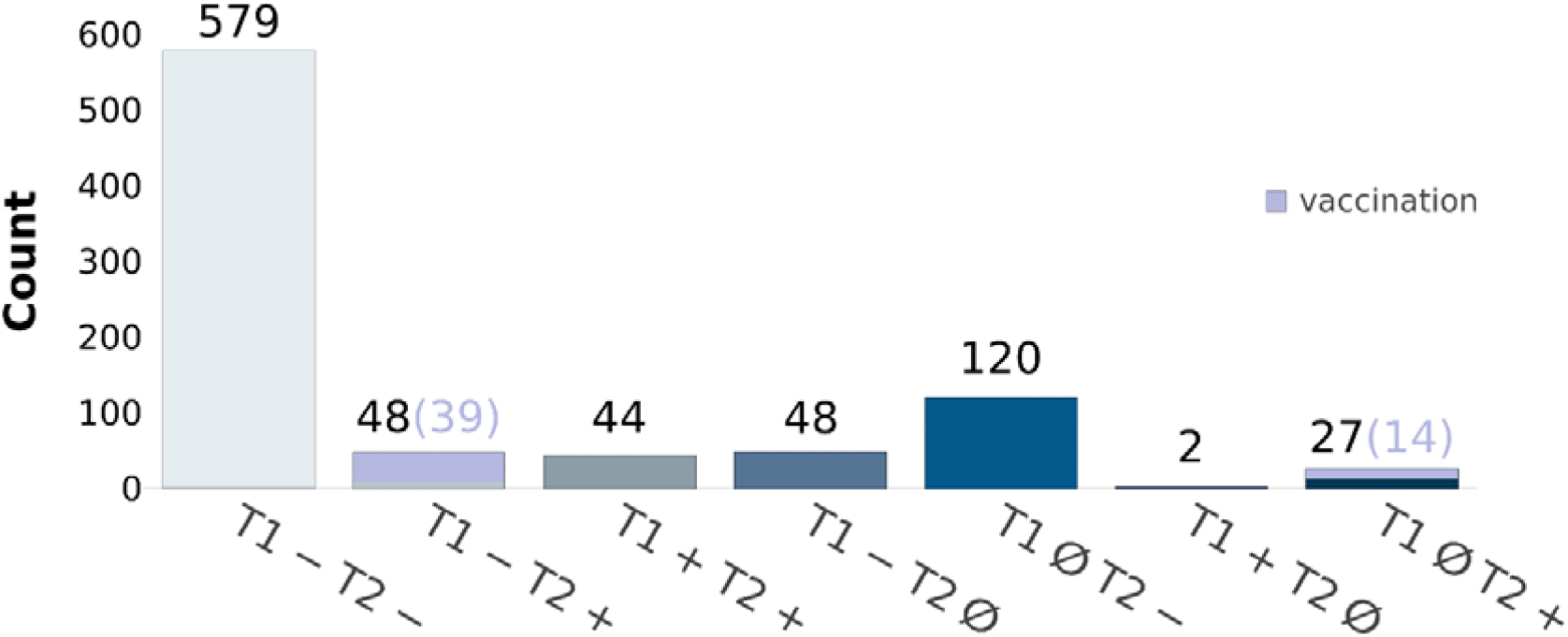
SARS-CoV-2 antibody status at the beginning and end of the semester across all cohorts Overall, eight of the seronegative individuals at the end of semester reported prior vaccination. However, either a complete report of vaccination was not available (dose, timing) or the initial dose was vaccinated a few days prior to blood collection. The number of vaccinated individuals with a positive result are indicated in brackets. T1 = antibody test semester start, T2 = antibody test semester end, Ø no participation.

**Supplementary Figure 4:**
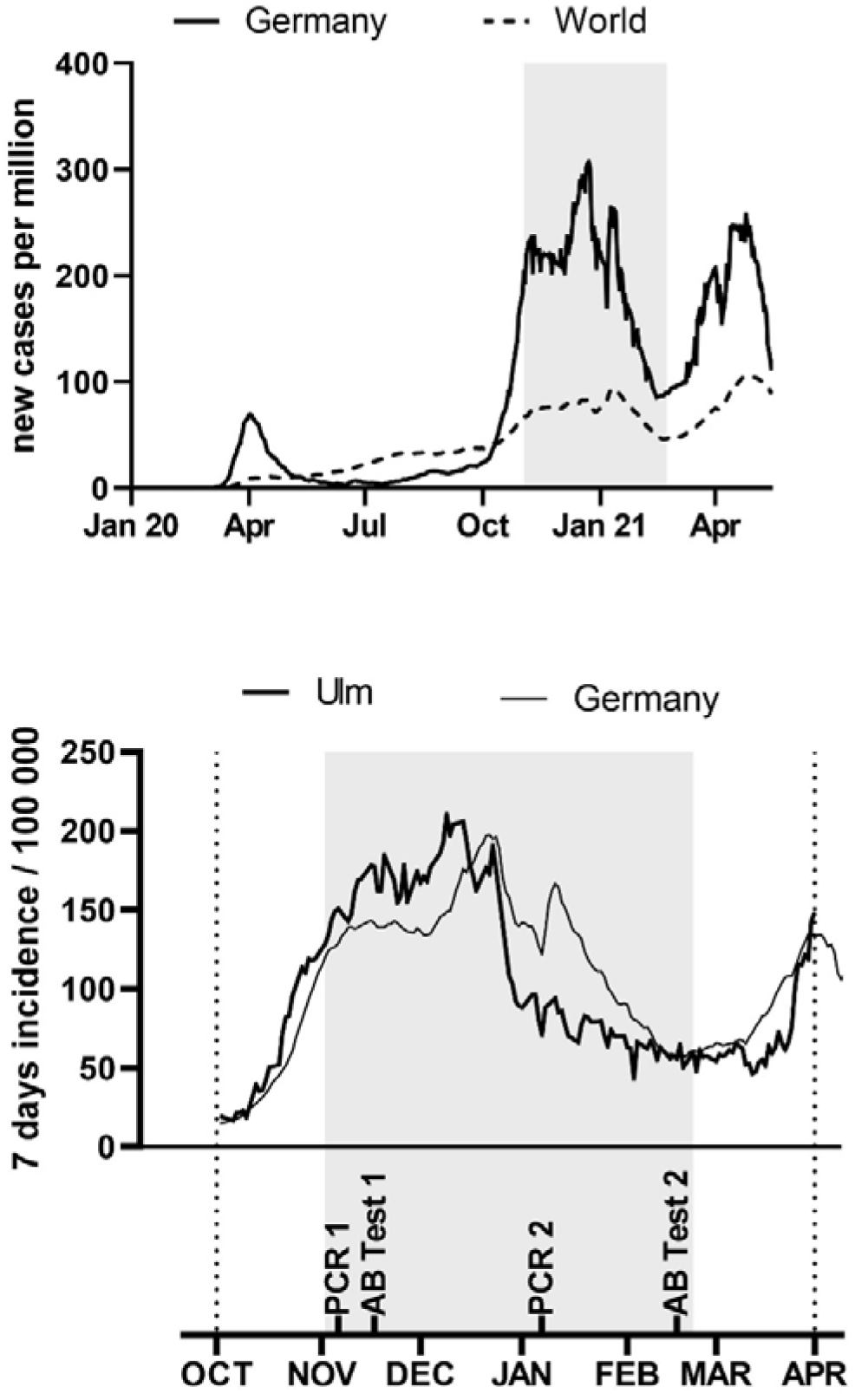
Worldwide, German, and city of Ulm incidence rates during the pandemic Top are the incidence rates of and Germany (continuous line) and worldwide (dashed line) during the pandemic shown (source for incidence rates: https://github.com/owid/covid-19-data/blob/master/public/data/owid-covid-data.csv). In the graph below, the period of the anatomical dissection course (gray area) is shown with incidence rates of Germany and the city of Ulm (source for incidence rates: Robert Koch Institute; https://www.rki.de/DE/Content/InfAZ/N/Neuartiges_Coronavirus/Daten/Fallzahlen_Kum_Tab.html). The incidences of infection in Ulm and Germany were very similar during the second wave. The test time points are plotted below.

**Supplementary Table 1:**
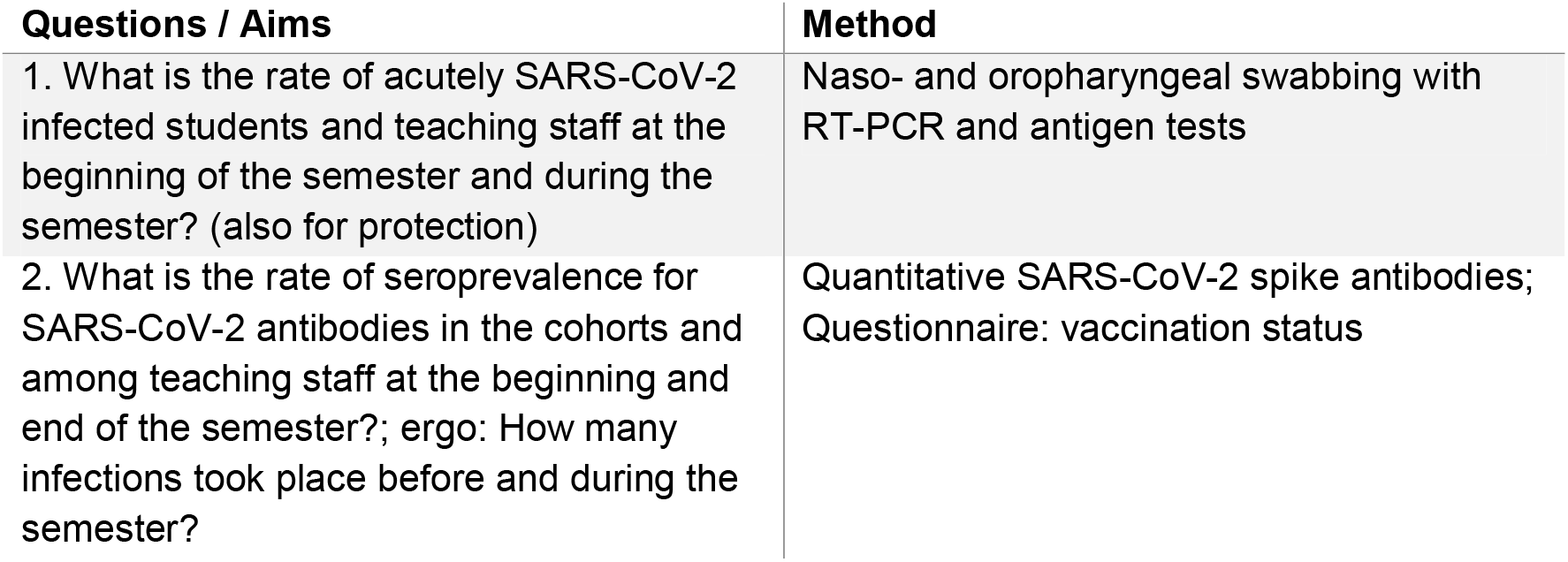
Aims and methods of the study

| Questions / Aims | Method |
| --- | --- |
| 1. What is the rate of acutely SARS-CoV-2 infected students and teaching staff at the beginning of the semester and during the semester? (also for protection) | Naso- and oropharyngeal swabbing with RT-PCR and antigen tests |
| 2. What is the rate of seroprevalence for SARS-CoV-2 antibodies in the cohorts and among teaching staff at the beginning and end of the semester?; ergo: How many infections took place before and during the semester? | Quantitative SARS-CoV-2 spike antibodies;<br>Questionnaire: vaccination status |

**Supplementary Table 2:** Hygiene concept for the gross anatomy course

| Measure | Explanation |
| --- | --- |
| spatial distance | - maximum of one person per 3.8 m<sup>2</sup><br>- maximum of six students and one tutor per table<br>- at least 1.5 m distance between table groups; timed entry and exit of table groups (also to relief public transport to university)<br>- oral examinations individually at the body donor<br>- written examinations with a minimum distance of 1.5 m between students |
| mask | - teaching staff: FFP2 masks<br>- students: FFP2 masks (at the beginning surgical mask) |
| hygiene | - constant wearing of medical gloves<br>- individualized protective coats (in the locker room in plastic envelopes); coats were regularly washed by the institute<br>- hand washing and disinfection when entering / leaving the course room<br>- daily disinfection of chairs and teaching aids such as models and computer pads |
| ventilation | - continuous supply of fresh air, complete air change approx. every 3 min, laminar air flow via the ceiling to openings near the floor |
| protection from infected individuals | - no participation in the course if symptoms suspicious for COVID-19 or if the person was a close contact (category I) according to the Robert Koch Institute |
| contact tracing | - attendance of each student was documented at the beginning of the course day<br>- additional tracing via a QR code-based contact tracing data app |
| information offer (also according to current university regulations) | - recurring information on hygiene rules, COVID-19 symptoms, behavior in case of infection, contact with infected persons, for risk groups, interim results of the study, and on the pandemic |

